# Symptoms of children and adolescents infected with SARS-CoV-2 variants alpha, delta or omicron

**DOI:** 10.1101/2024.01.07.23300006

**Authors:** Hansjörg Schulze, Wibke Bayer

**Affiliations:** Institute for Virology, University Hospital Essen, University Duisburg-Essen, Essen, Germany

**Keywords:** SARS-CoV-2, symptoms, children, adolescents, mild disease

## Abstract

Symptoms experienced by children and adolescents with SARS-CoV-2 infections in the alpha, delta and omicron variant dominated phases were investigated using an online survey, and the frequencies of reported symptoms and changes over time were analyzed. The most prevalent symptoms were fever above 38 °C, tiredness, headache, runny or blocked nose, sneezing and dry cough. Lethargy and nausea were reported significantly more frequently in the omicron variant dominated phase than in the earlier phases of the pandemic. Compared to symptoms reported by adults, fever and gastrointestinal symptoms were reported more frequently for children, especially in the omicron variant dominated phase, whereas the frequency of loss of smell and loss of taste was significantly lower in children than in adults.

## Introduction

The SARS-CoV-2 pandemic started in late 2019 in Wuhan, China, where the first cases and human-to-human transmission were reported [1], and the virus spread quickly all over the world. By August 2023, more than 769 million people have been infected according to data from the World Health Organization [2], which is very likely an underestimate since testing for SARS-CoV-2 has been reduced significantly since spring 2023 as SARS-CoV-2 has no longer been considered a public health emergency [3].

SARS-CoV-2 can induce severe disease in adults as well as in children, but mild disease is most common in both children and adults. The rate of asymptomatic infections is generally hard to determine and reported numbers vary widely; different meta-analyses calculated asymptomatic infection rates in children in the earlier phases of the pandemic of 19.3% [4], 13.1% [5] or 46.7% [6]. In the symptomatic infections, cough, nausea, vomiting and diarrhea have been reported as the most frequent symptoms of children during the early, SARS-CoV-2 D614G dominated phase of the pandemic [5]. For later, variant-dominated phases, increases in the frequency of fever, cough and upper respiratory tract symptoms have been described [7].

We have previously reported on a detailed study of self-reported symptoms by SARS-CoV-2 infected adults during four different phases of the pandemic, where we determined the frequency of more than 40 different symptoms and analyzed changes in the variant-dominated phases [8]. Our new data now provide detailed insight into symptoms experienced by SARS-CoV-2 infected children in phases of the pandemic that were dominated by different SARS-CoV-2 variants, and we present a side-by-side comparison with symptom frequencies reported by adults.

## Methods

### Ethics Approval

The study was approved by the local Ethics Committee of the Medical Faculty of the University Duisburg-Essen (approval number 20-9233-BO). The study was carried out in accordance with the ethical guidelines and regulations. The study participants and a parent or another legal representative gave their informed consent to their voluntary participation in the study and to the subsequent use of the data for publication.

### Data collection

Data were obtained using an online questionnaire based on the LimeSurvey software which was hosted on the servers of the University Duisburg-Essen. Participants for the study were recruited via the public health offices of the administrative district of Soest (North Rhine-Westphalia, Germany) and of the administrative district Hochsauerlandkreis (North Rhine-Westphalia, Germany) when they received their confirmation of a positive SARS-CoV-2 PCR test result. The data presented in this manuscript were collected between January 23^rd^ 2021 and March 22^nd^ 2022. During this time, official SARS-CoV-2 antigen testing was accessible in Germany when a SARS-CoV-2 infection was suspected with or without respiratory symptoms or prior contact. Semi-weekly self-testing or pool-testing was introduced from April 2021 for children visiting a kindergarten or school. An individual PCR test was mandatory after a positive official or at-home antigen test or pool PCR test. Official PCR test results and resulting quarantines were managed by the public health offices.

### Data analysis

Data were analyzed using R (version 3.6.3) and RStudio software and fmsb, plyr, splyr, tidyverse, reshape2, stats, ggplot2 and viridis packages. Only data from individuals who had completed the survey were included in the analysis.

For graphical representation, a heatmap of the data was created with ggplot2 after sorting the responses by the date of survey completion and by the age of the respondents. The category “fever > 38.0 °C” was added after calculation from the survey responses using the tidyverse package.

The test for statistically significant differences in symptom frequencies between SARS-CoV-2 infected children in the alpha, delta and omicron variant dominated phases and in symptom frequencies between children from the current survey and adults from our previous symptom study [8] was performed using Fisher’s exact test using the function fisher.test of the R stats package. The test for statistically significant differences in the age distribution of the respondents from the alpha, delta and omicron variant dominated phases was performed by One Way analysis of variance (One Way ANOVA) using the function aov() of the R stats package.

## Results

An online based survey was performed to gather information on the prevalence of different symptoms in children and adolescents after infection with SARS-CoV-2. Participants were recruited by their local health office. In total, 101 responses were obtained from or for children and adolescents aged between 0 years and 18 years (Table 1). The study started in January 2021 during the SARS-CoV-2 alpha variant dominated phase (in Germany: February 2021 – June 2021), and data were collected over a period of more than a year, thus also covering the delta variant (July 2021 – January 2022) and omicron variant dominated phases (from January 2022; [9]). The major characteristics of the participants from the three phases are comparable, with a majority of respondents between 7 and 12 years old. The subjective assessment of the severity of the infection, ranging from asymptomatic over mild (e.g.: “symptoms without feeling ill”) and moderately ill (e.g.: “feeling ill and lying on sofa”) to very ill (e.g.: “feeling ill and having to stay in bed”) resulted in a majority (overall 50.5%, alpha: 58.3%, delta: 43.2%, omicron: 55.6%) of respondents answering with “moderately ill”. Only a minority of 5.9% overall (alpha: 8.3%, delta 11.4%, omicron 0%) reported an asymptomatic infection. None of the respondents reported a hospitalization or the necessity of intensive care treatment due to the SARS-CoV-2 infection (0%).

**Table 1.** Characteristics of the study population.

| subgroup number | all respondents 101 | alpha 12 | delta 44 | omicron 45 |
| --- | --- | --- | --- | --- |
| **age (years)** |  |  |  |  |
| 0-6 | 27 (26.7%) | 3 (25%) | 14 (31.8%) | 10 (22.2%) |
| 7-12 | 61 (60.4%) | 6 (50%) | 26 (59.1%) | 29 (64.4%) |
| 13-18 | 13 (12.9%) | 3 (25%) | 4 (9.1%) | 6 (13.3%) |
| **gender** |  |  |  |  |
| female | 54 (53.5%) | 4 (33.3%) | 26 (59.1%) | 24 (53.3%) |
| male | 47 (46.5%) | 8 (66.6%) | 18 (40.9%) | 21 (46.7%) |
| **smoking** |  |  |  |  |
| active | 0 (0%) | 0 (0%) | 0 (0%) | 0 (0%) |
| passive | 6 (5.9%) | 1 (8.3%) | 2 (4.5%) | 3 (6.7%) |
| **hayfever<sup>1</sup>** | 4 (3.9%) | 0 (0%) | 1 (2.3%) | 3 (6.7%) |
| **severity** |  |  |  |  |
| asymptomatic | 6 (5.9%) | 1 (8.3%) | 5 (11.4%) | 0 (0%) |
| mild<sup>2</sup> | 34 (33.7%) | 3 (25%) | 19 (43.2%) | 12 (26.7%) |
| moderately ill<sup>3</sup> | 51 (50.5%) | 7 (58.3%) | 19 (43.2%) | 25 (55.6%) |
| very ill<sup>4</sup> | 10 (9.9%) | 1 (8.3%) | 1 (2.3%) | 8 (17.8%) |
| hospitalization | 0 (0%) | 0 (0%) | 0 (0%) | 0 (0%) |
| intensive care treatment | 0 (0%) | 0 (0%) | 0 (0%) | 0 (0%) |

<sup>1</sup> hayfever relevant at the time of infection
<sup>2</sup> e.g. “I did not feel ill but had a runny nose”
<sup>3</sup> e.g. “I felt ill and lay on the sofa”
<sup>4</sup> e.g. “I felt very ill and had to stay in bed”

The symptoms reported by the survey respondents are shown in Figure 1A as a heatmap and as bargraphs in Figure 2. The most frequently reported symptoms included general symptoms such as fever, tiredness and headaches, and more specific respiratory symptoms such as a runny or blocked nose and dry cough. The comparison of the symptoms reported during the three different phases of the pandemic (Figure 2B), i.e. the alpha, delta, and omicron variant dominated phases, revealed only minor differences in symptom frequencies with the exception of blocked nose, which dropped from 83.3% in the alpha variant dominated phase to 47.7% and 42.2.% during the delta and omicron variant dominated phases, respectively, and changes in the frequency of lethargy (alpha: 25%, delta: 22.7%, omicron: 53.3%), loss of smell (alpha: 8.3%, delta: 18.2%, omicron: 4.4%) and nausea (alpha: 16.7%, delta: 9.1%, omicron: 33.3%). Importantly, differences observed in the three phases do not correlate to a difference in the age distribution of the respondents (Figure 1B, no significant differences, One Way ANOVA).

**Figure 1.**
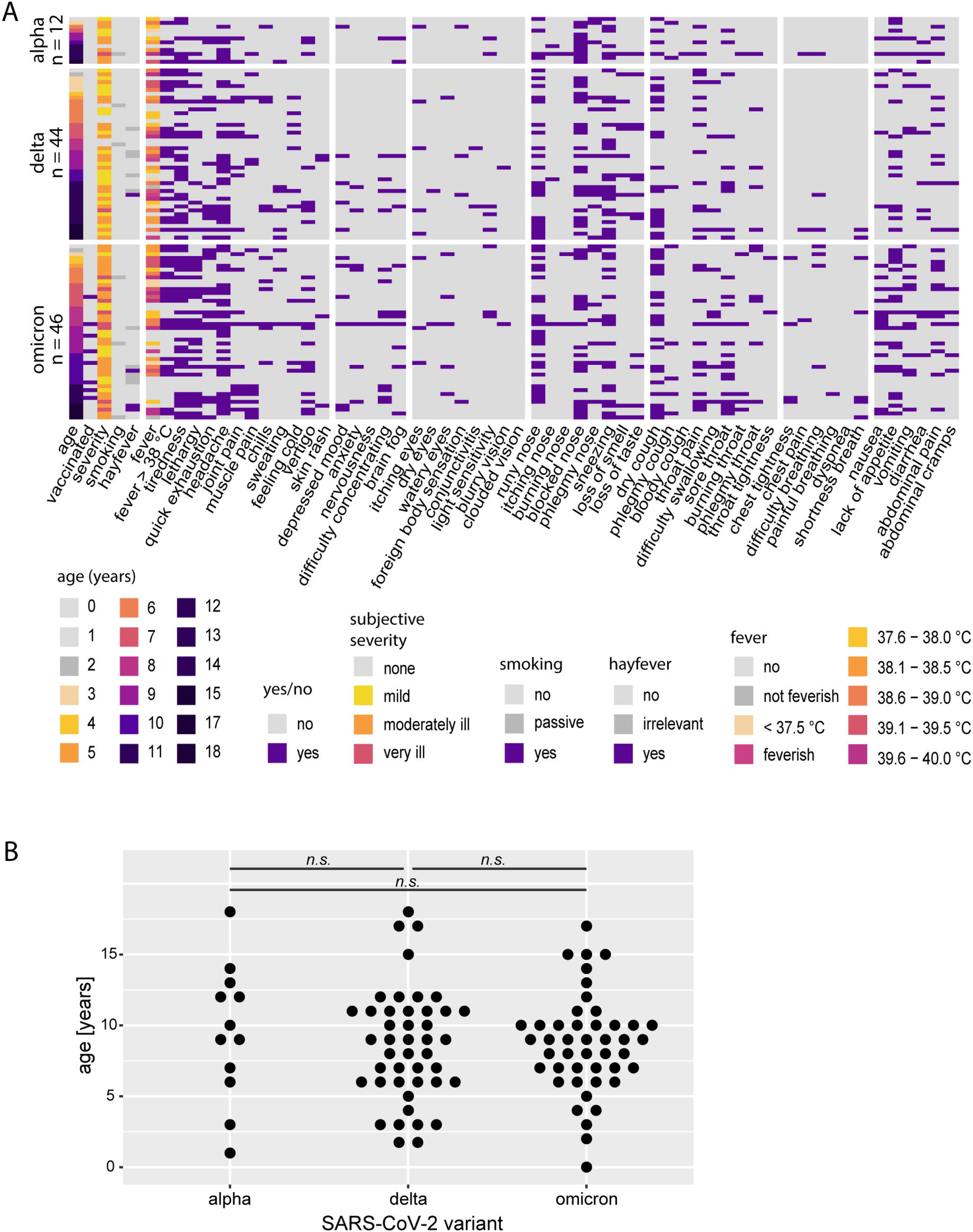
Heatmap of symptoms of SARS-CoV-2-infected children and adolescents. (A) Symptoms of SARS-CoV-2 infected children and adolescents as reported during the alpha (top), delta (delta) and omicron dominated phases (bottom) of the SARS-CoV-2 pandemic, responses were sorted by variant phase and age of the participant. “feverish/not feverish”: subjective judgement, body temperature was not measured. (B) The graph shows the age of the respondents from the alpha, delta and omicron variant dominated phases of the SARS-CoV-2 pandemic. Each dot indicates an individual, n.s. = not significant (*P* > 0.05, One Way ANOVA).

**Figure 2.**
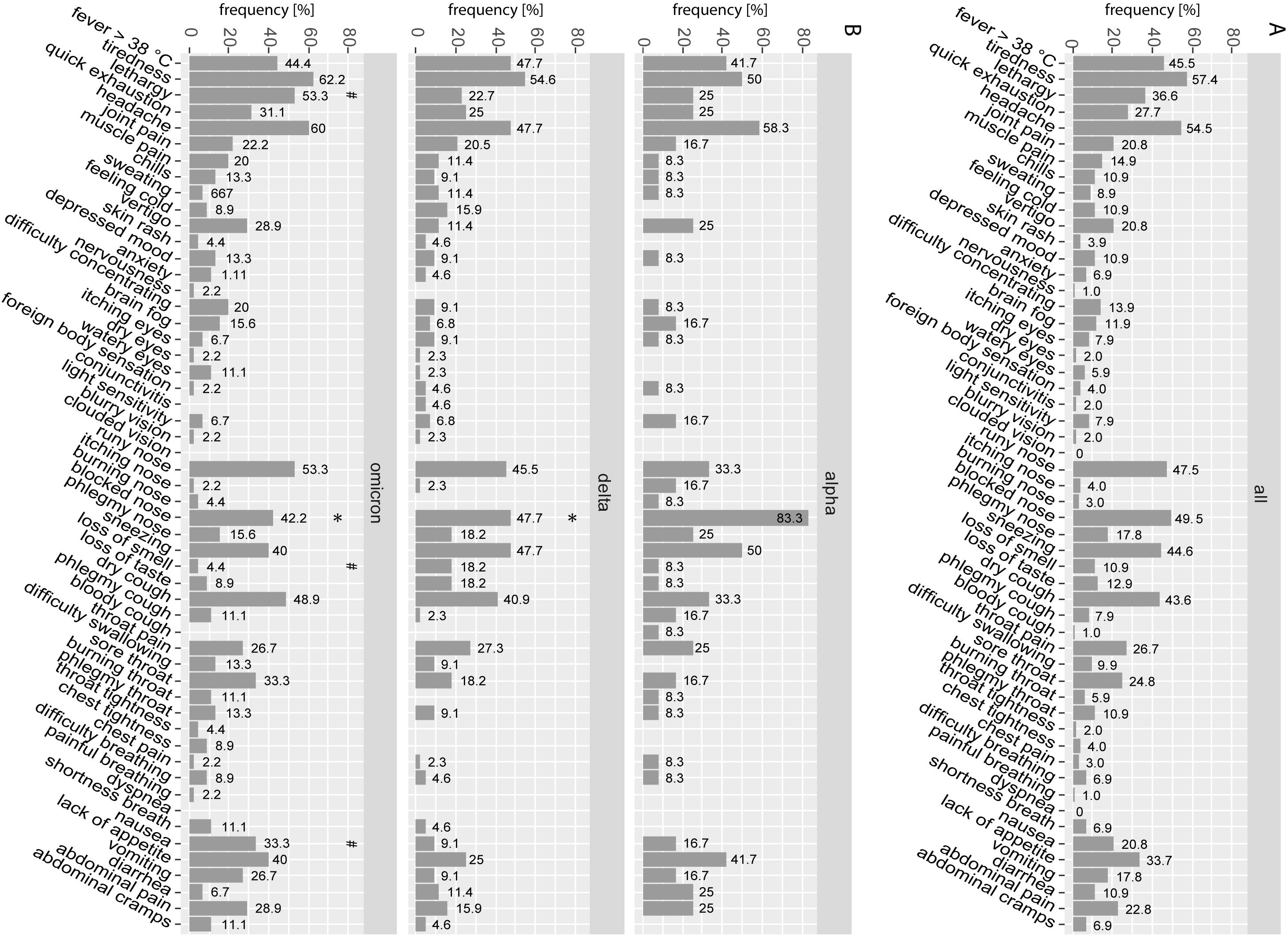
Frequency of symptoms stratified by SARS-CoV-2 variant. Barplots indicate the frequency of the indicated symptoms reported by all respondents (A) or by the respondents infected during the alpha, delta or omicron dominated phases (B). * indicates statistically significant differences compared to the frequency during the alpha variant dominated phase, # indicates statistically significant differences compared to the frequency during the delta variant dominated phase (*P* < 0.05, Fisher’s exact test).

We have reported before on the results of a similar symptom survey where we gathered information about symptoms experienced by adults infected with SARS-CoV-2 [8]. Performing a side-by-side comparison, we found that symptom frequencies during all three variant dominated phases differed between adults and children, with most general symptoms such as lethargy (alpha: adults 60.5%, children 25%; delta: adults 50.2%, children 22.7%; (omicron: adults 46.5%, children 53.3%)), headache (alpha: adults 68.3%, children 58.3%; delta: adults 67.4%, children 47.7%; omicron: adults 65.7%, children 60%), muscle pains (alpha: adults 31%, children 8.3%; delta: adults 28.4%, children 11.4%; omicron: adults 30.4%, children 20%) and sweating (alpha: adults 21.9%, children 8.3%; delta: adults 28.4%, children 11.4%; omicron: adults 21%, children 6.7%) reported at significantly lower frequency for SARS-CoV-2 infected children compared to SARS-CoV-2 infected adults (Figure 3). While the frequencies of the hallmark symptoms of SARS-CoV-2 infection, loss of smell and taste, were also significantly lower in children in all three variant dominated phases (loss of smell/taste, alpha: adults 48.2%/42%, children 8.3%/8.3%; delta: adults 60.6%/54.6%, children 18.2%/18.2%; omicron: adults 25%/23.8%, children 4.4%/8.9%), other symptom frequencies were elevated in children compared to adults, notably the frequency of fever higher than 38 °C (alpha: adults 19.4%, children 41.7%; delta: adults 17.9%, children 47.7%; omicron: adults 16.6%, children 44.4%) and gastrointestinal symptoms such as nausea (alpha: adults 19.4%, children 16.7%; delta: adults 12.9%, children 9.1%;) omicron: adults 11.7%, children 33.3%), vomiting (alpha: adults 2.7%, children 16.7%; delta: adults 1.8%, children 9.1%; omicron: adults 1.2%, children 26.7%), abdominal pain (alpha: adults 10.3%, children 25%; delta: adults 8.5%, children 15.9%; omicron: adults 8.9%, children 28.9%) and abdominal cramps (alpha: adults 6.5%, children 0%; delta: adults 5%, children 4.5%;) omicron: adults 3.5%, children 11.1%).

**Figure 3.**
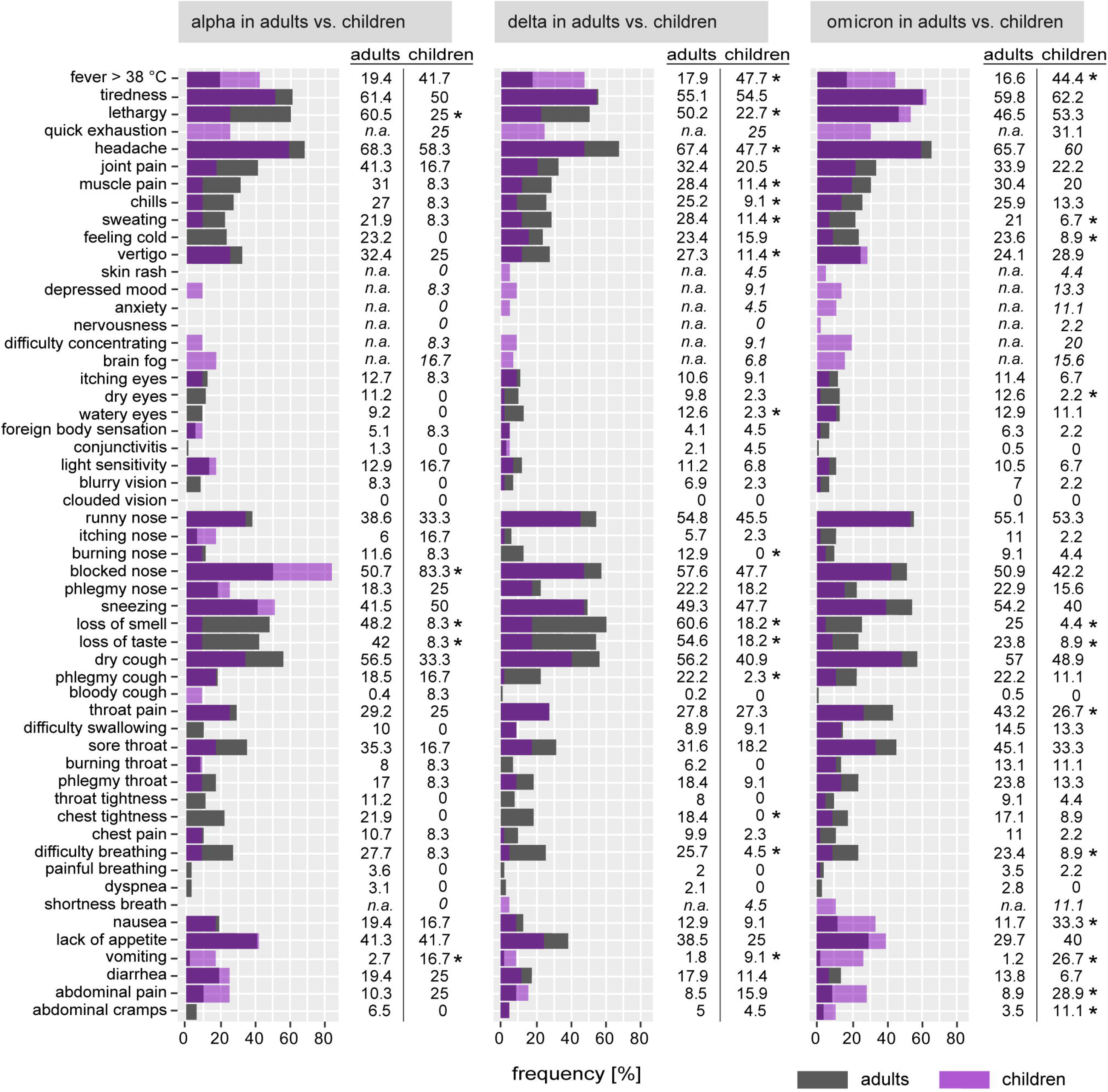
Comparison of symptom frequencies reported by adults and children. The frequencies of symptoms reported by adults have been reported before in a previous publication [8] and are shown by grey bars. The frequencies of children are shown as purple bars. Significant differences between frequencies reported by adults and children are indicated by * (*P* < 0.05, Fisher’s exact test). n.a. = not available: data were not acquired in the previous study.

## Discussion

In this report, we provide a detailed analysis of the symptoms experienced by SARS-CoV-2 infected children, and show that the symptoms reported most often and at rates of more than 40% were fever above 38 °C, tiredness, headache, runny nose, blocked nose, sneezing and dry cough. It is interesting to note that in comparison to symptom frequencies reported by adults, we observed many minor differences, and a significantly higher frequency of fever and a strikingly lower frequency of loss of smell and taste was observed in children compared to adults.

It has been reported before that a very high percentage of SARS-CoV-2 infected children experienced fever, and rates in other reports are often even higher than the frequency reported by us [4; 5; 7]. We found that the frequency of fever above 38 °C was significantly higher in children compared to adults, which may be explained by the finding that the expression levels of type I interferons are higher in children than in adults [10; 11; 12; 13; 14].

The loss of smell and/or loss of taste has been considered a hallmark symptom of SARS-CoV-2 infection since the beginning of the pandemic [15], and was shown by us [8] and others to have decreased in frequency in adults infected with SARS-CoV-2 since the spread of the SARS-CoV-2 omicron variant [16] from a frequency around 60% to below 20%. The frequency in children however was much lower in all three phases of the pandemic that we analyzed here, which is in agreement with findings by others; it was shown in a meta-analysis that the frequencies in individual studies varied markedly and that children of more than ten years of age were more likely to experience loss of smell or taste [17].

There are some limitations to our study, most importantly that the total number of respondents was low, and children below the age of 6 and between 13 and 18 years old are less represented than the age group of 7 to 12 years old. We were therefore not able to correlate symptom frequencies with age groups, and would also like to point out the relatively low number of respondents with regard to the comparison of symptom frequencies for the three different SARS-CoV-2 variants. Furthermore, the symptoms of SARS-CoV-2 infected children were either self-reported or reported by a parent and can be subjective, and the presence of some symptoms such as headache, joint pain or muscle pain can be difficult or impossible to assess or to distinguish in very young children. As is always the case with self-report questionnaires, there is a certain response bias in that it is more likely that infected individuals with symptoms, or in this case the parents of the young individuals, take part in the study than asymptomatic individuals. Even though a few individuals reported no symptoms, this rate should not be construed as the rate of asymptomatic infection. In many cases, the questionnaires were completed within a week after the positive PCR test, but in general the study has to be considered partly retrospective, which may have resulted in a less clear picture than a daily questionnaire. With regard to the attribution of the infections to a certain SARS-CoV-2 dominated phase, it has to be pointed out that infection with a certain variant was not confirmed and that the attribution was performed solely on the date of questionnaire completion. While the alpha, delta and omicron variants quickly replaced the respective preceding variants [9], there were short times of overlap when both variants were present, which slightly weakens the allocation of responses to the variants. Finally, we did not obtain information in this study about the duration of symptoms, and cannot assess how long individual symptoms persisted.

Overall, our study provides additional insight into the symptoms experienced by SARS-CoV-2 infected children and adolescents and contributes to understanding this infection in the young.

## Ethics Statement

The studies involving human participants were reviewed and approved by Ethik-Kommission der Medizinischen Fakultät der Universität Duisburg-Essen Robert-Koch-Str. 9-11 45147 Essen Germany. The patients/participants provided their informed consent to participate in this study.

## Author Contributions

HS recruited participants and contributed to writing the manuscript. WB conceived of the study, created the online survey, analyzed data and wrote the manuscript. All authors contributed to the article and approved the submitted version.

## Acknowledgments

We would like to thank all participants for taking part in our survey.

## Data Availability Statement

The raw data supporting the conclusions of this article will be made available by the authors, without undue reservation.

